# Insights into *DEPDC5*-Related Epilepsy from 586 people: Variant Penetrance, Phenotypic Spectrum, and Treatment Outcomes

**DOI:** 10.1101/2024.12.25.24319647

**Authors:** Manuela Ochoa-Urrea, Elizabeth A. Butler, Tobias Bruenger, Costin Leu, Christian M. Bosselmann, Imad Najm, Samden Lhatoo, Dennis Lal

## Abstract

Variants in the GATOR1 complex gene *DEPDC5* disrupt mTORC1 pathway regulation, driving cortical malformations and focal epilepsy. *DEPDC5* is the most common genetic cause of focal epilepsy, often linked to focal cortical dysplasia (FCD). However, reduced penetrance complicates genetic counselling and risk prediction.

This study analysed the largest *DEPDC5*-related epilepsy cohort, synthesising data from 170 families and 586 variant carriers. Epilepsy penetrance was estimated at 64.9% (*n=*380/586), with median seizure onset at 5 years. By age 10, 76.1% of individuals experienced seizures. Drug resistance occurred in 48.3% (*n=*101/209) of cases, and cortical malformations were present in 28% (*n=* 49/175) of MRIs. SUDEP accounted for 16% (*n=* 4/25) of deaths among affected individuals.

Early seizure onset correlated strongly with drug resistance, intellectual disability, and MRI abnormalities, underscoring its role as a severity marker. Surgical intervention in drug-resistant cases (34.7% [*n=* 35/101]) achieved favourable outcomes (Engel I or II) in 88% (*n=* 29/33) of individuals, with pathology confirming cortical malformations in 92.6% (*n=*25/27) of those with abnormal MRIs.

This study advances the understanding of *DEPDC5*-related epilepsy, providing critical insights into penetrance, phenotype, and treatment outcomes to inform precision care and genetic counselling.

## Introduction: Understanding *DEPDC5*-related epilepsy

DEP domain containing 5 (*DEPDC5*) gene encodes for a key component of the GTPase□activating protein activity toward Rags 1 (GATOR1) protein complex. The GATOR1 complex regulates the mechanistic target of the rapamycin complex 1 (mTORC1) pathway by inhibiting its activity when amino acids are in low supply.^1^ In the developing brain, proper mTORC1 signalling is critical for normal neural proliferation, growth and differentiation, and circuit formation.^1–3^ Pathogenic variants in *DEPDC5* gene result in a loss of GATOR1 function, leading to hyperactivation of mTORC1 signalling in the brain, independent of amino acid availability.^2^ This overactivity disrupts normal brain development, alters cortical structures, and may increase neuronal excitability.^2^

Pathogenic germline variants in *DEPDC5* were first implicated in focal epilepsy through exome sequencing of families with autosomal dominant familial focal epilepsies with variable foci.^4,5^ Since then, *DEPDC5* variants have now been associated with a broad range of epilepsies, including infantile spasms, self-limited epilepsy with centrotemporal spikes, structural epilepsy secondary to focal cortical dysplasia (FCD) and hemimegalencephaly.^4–13^. Pathogenic *DEPDC5* variants cause 10% of focal epilepsy cases, making them the most common genetic cause.^13^ The overall incidence of *DEPDC5*-related epilepsies has been estimated to be 2.36 per 100,000 births (90% CI: 0.81-5.59).^14^ Their relatively high frequency and high hereditability stem from factors like reduced penetrance—meaning not everyone with the variant will show symptoms—and often less severe epilepsy phenotypes. Prior family studies have estimated that approximately 66% (*n*= 69/105, 7 families) to 69% (*n*= 231/335, 94 families) of variant carriers will develop epilepsy.^5,15^

The clinical presentation of *DEPDC5* variant carriers displays considerable heterogeneity, with variability observed among family members.^13^ In an international retrospective cohort study of 63 probands, affected individuals typically presented with childhood-onset focal seizures, one-third of whom will present within the first year of life.^13^ Seizures occurred during sleep in 48% of the individuals. Thirty-five per cent of the seizures were classified as hyperkinetic, and 10% presented with infantile epileptic spasms.^13^ Drug resistance was reported in 32/61 of this cohort, 25% of whom underwent epilepsy surgery with good outcomes.^13^ Intellectual disability, usually mild, was identified in 27% of affected individuals, and psychiatric comorbidities—such as attention deficits, oppositional behaviours, mood disorders, and autism—affected 46% of the studied cohort.^13^ Sudden unexplained death in epilepsy (SUDEP) occurred in 9-11% of the families, underscoring the importance of vigilant clinical management.^13^

Previous cohort studies on *DEPDC5*-related epilepsy have struggled to establish clear genotype-phenotype correlations due to small cohort sizes, inconsistent reporting of key clinical features (such as drug resistance, MRI findings, intellectual disability, and psychiatric comorbidities), and a predominance of participants of European ancestry. This narrow scope has made it difficult to determine how *DEPCD5* variants influence disease severity, clinical course, or penetrance across different ethnic backgrounds. As a result, providing accurate risk stratification and identifying reliable biomarkers for treatment outcomes has been challenging. The natural history and prognosis of *DEPDC5*-related epilepsy, particularly in cases with early seizure onset, remain poorly understood, complicating risk counselling and family planning. While preclinical and observational data suggest that mTOR inhibitors may be a promising treatment,^16–18^ clinical trials are needed to confirm their efficacy and establish prognostic markers for precision medicine. Recent cohort studies involving underrepresented populations, including several from Asian countries, have begun to enhance the diversity of studied cohorts and improve the applicability of findings across different ethnic groups. However, significant gaps remain, especially concerning genotype-phenotype correlations, disease natural history, and penetrance in diverse populations.

To address these gaps, we comprehensively synthesised data from 33 studies involving 170 families and 586 *DEPDC5* variant carriers. This pooled analysis offers an expanded view of the clinical spectrum of familial *DEPDC5*-related epilepsy, identifies novel clinical associations, and improves penetrance estimates to refine risk assessment and genetic counselling. Our findings aim to inform clinical decision-making, support family planning, and advance personalised treatment strategies while laying the groundwork for future research into targeted therapeutic approaches.

## Materials and methods

### Publication search strategy

A scoping literature review was conducted following the PRISMA-ScR framework (Preferred Reporting Items for Systematic Reviews and Meta-Analyses Extension for Scoping Reviews), to ensure methodological rigor and reproducibility (Supplementary Table 1).^19^ All studies listed on PubMed until August 31, 2024 were considered, using the gene-specific term “*DEPDC5*” as the keyword for the search. Two investigators reviewed the publications in two steps: first, a title and abstract screening excluded non-English, non-neurological, or non-genetic studies. Second, a full-text review selected studies with genotype and phenotype data from families meeting specific criteria, excluding individual cases, sporadic cases, and recessive inheritance reports; duplicate families were removed by cross-referencing citations, pedigrees, and phenotype details. For a detailed description of the process, see supplementary methods.

### Data collection and organisation

We extracted genotype and phenotype information from 33 publications identified in the literature review (details in Supplementary Methods and Supplementary Table 2). Data collected included the type of *DEPDC5* variant, epilepsy phenotype, and various clinical variables such as seizure onset age, epilepsy type, drug resistance, neuroimaging results, comorbidities, epilepsy surgery, and SUDEP. Obligate carriers—identified through pedigree analysis and reported clinical data—were included to capture the inheritance dynamics of *DEPDC5* variants. Additional clinical details were gathered from supplementary reports (Supplementary Table 3). To examine inheritance patterns, a sub-cohort of 68 multiplex families (families in which multiple individuals are affected by epilepsy)^20^ with confirmed pathogenic *DEPDC5* variants and epilepsy phenotypes was analysed, excluding families with limited sequencing data. Full methodological details are provided in the Supplementary Methods section.

### Statistical analysis and penetrance calculation

All statistical analyses were conducted using R (version 4.4.0)^21^ with clinical data from individuals with confirmed or inferred *DEPDC5* variants. Fisher’s Exact Test was used to evaluate the enrichment of clinical features in drug-resistant epilepsy cases, and the Wilcoxon Rank Sum Test assessed associations between the age of seizure onset and clinical variables. The nominal significance level was set at α = 0.05. Cumulative seizure onset incidence and 95% confidence intervals were calculated from standard error estimates. Penetrance was determined as the proportion of heterozygous *DEPDC5* variant carriers with an epilepsy phenotype and compared between multiplex families and all families using a two-tailed t-test. Detailed methods are provided in the Supplementary Methods section.

## Results

### Literature review and penetrance results

Here, we present a scoping review of 226 unique studies. The abstract and title were reviewed for all (see Figure 1 for the selection process diagram). Ultimately, 33 studies were included in our analysis, representing 170 unique families and 586 individuals with confirmed or obligate carriers of *DEPDC5* variants. We extracted and curated clinical and genetic data for 96.1% (*n=*563/586) confirmed heterozygous variant carriers and 3.9% (*n=*23/586) assumed carriers. Across all 170 families, 380/586 individuals had a diagnosis of epilepsy, leading to an estimated *DEPDC5*-related epilepsy penetrance of 64.9% (95% CI: 60.8-68.7%). Of those with *DEPDC5*-related epilepsy, 98.2% (*n=*373/380) carried a verified variant, and 1.8 (*n=*7/380) were inferred carriers.

**Figure 1.**
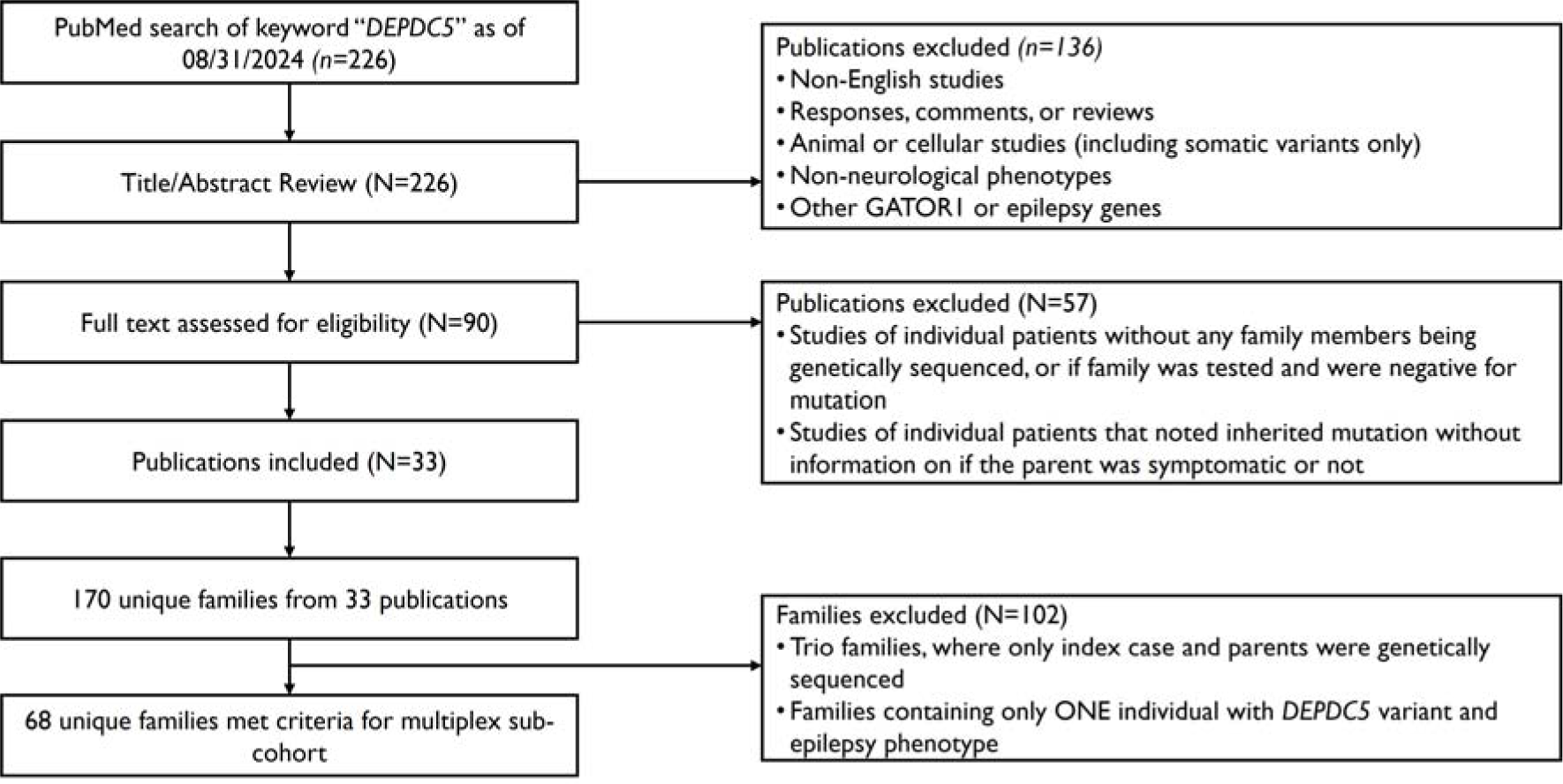
PRISMA flow diagram. Review screening process and family cohort filtering.

Sixty-eight multiplex families were identified, comprising 264 individuals with epilepsy out of 375 heterozygous variant carriers or obligate carriers, with an estimated penetrance for epilepsy in multiplex families of 70.4% (95% CI: 65.5-75.0%). We observed no significant difference in mean penetrance for epilepsy when compared with the estimated epilepsy penetrance from the multiplex subgroup (two-tailed t-test: *P*=0.51)

### Description of clinical features

Clinical characteristics were analysed in individuals with confirmed and inferred *DEPDC5* variants and epilepsy. The median age of onset of epilepsy was five years (IQR 1-10; Table 1). By the age of 1 year, 26.9% of individuals had developed epilepsy, and at age 5 years, 51% had experienced their first seizure. By 10 years of age, 76.1% of the individuals had developed epilepsy (Figure 2). Focal epilepsy was the most common epilepsy type (83.9% [*n=* 319/380]). Among individuals with available MRI data, 33.1% showed lesions (Figure 2B and 2C), predominantly malformations of cortical development (MCD) (85.9% [*n=* 49/58] of all reported lesions). Hemimegalencephaly was reported in one individual without a familial history of epilepsy.^13^ No tubers or hamartomas were reported. Drug resistance was reported in 48.3% (*n=*101/209) of the individuals (Figure 2C), and epilepsy surgery was performed in 35% (*n=*35/101) of these individuals. Surgery outcomes were favourable in most cases, with 81.8% (27/33) achieving Engel I (free from disabling seizures) and 6% (2/33) Engel II (rare disabling seizures). The proportion of individuals who attained good surgical outcomes was similar between individuals who experienced epilepsy onset before or at age 5 years (*n*=28, Engel I or II: 82.14%) and those who had later seizure onset (*n*=6, Engel I or II: 100%). Individuals with MCD detected on MRI received surgical intervention 18.1-fold more frequently than those with reported normal MRI (61.3% vs 7.84%; Fisher’s Exact Test, 95%CI [6.7-54.5], *P* = 2.5e-11), and pathology confirmed MCD in 92.6% of the cases. The most frequent histopathology was FCD IIa, (60.6% [*n=*20/33]) (Table 1). Of individuals who received surgery and had non-lesional MRIs, 71.43% (*n=*5/7) were found to have FCD on the pathology specimen. Among individuals with MRI-detected MCD, 80.7% (*n=*21/26) achieved a surgical outcome of Engel I compared to 85.7% (*n=*6/7) individuals with non-lesional MRI.

**Figure 2.**
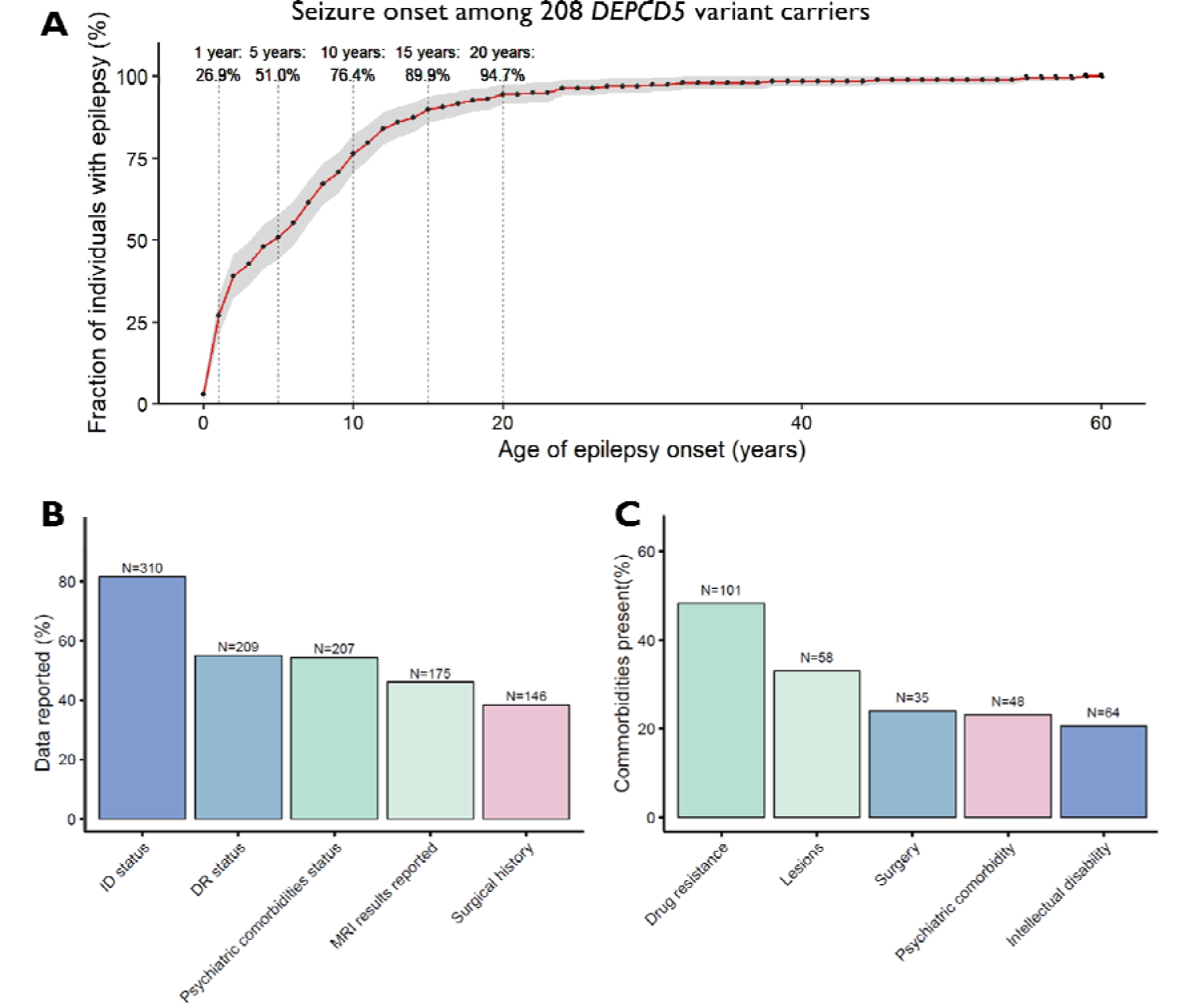
Key clinical features. **A)** Calculated cumulative incidence of epilepsy for specific age ranges across all variant carriers with *DEPDC5*-related epilepsy. The graph shows the cumulative fraction of individuals (y-axis) who experienced epilepsy onset by a given age (x-axis) across 208 individuals with reported ages of seizure onset. For example, 26.9% of individuals had epilepsy onset by the age of 1 year, and by 10 years, 76.4% of *DEPDC5* variant carriers with epilepsy had experienced their first seizure. The cumulative incidence of age of seizure onset was calculated by determining the proportion of individuals who had experienced their first seizures at each age, derived exclusively from the group of individuals with *DEPDC5*-related epilepsy, excluding variant carriers without epilepsy. B) Bar graph representing the percentage of affected individuals (out of *n=*380) with available clinical data for intellectual disability (ID), psychiatric comorbidities, drug resistance (DR), lesions, and surgical history. C) For those with available data on specific clinical features, the bar graph depicts the percentage of individuals exhibiting each comorbidity. For instance, of the 209 individuals with data on drug resistance (Figure 2B), approximately 48% (*n=*101, Figure 2C) were reported as drug-resistant. ID; intellectual disability; DR; drug resistance.

**Table 1.**
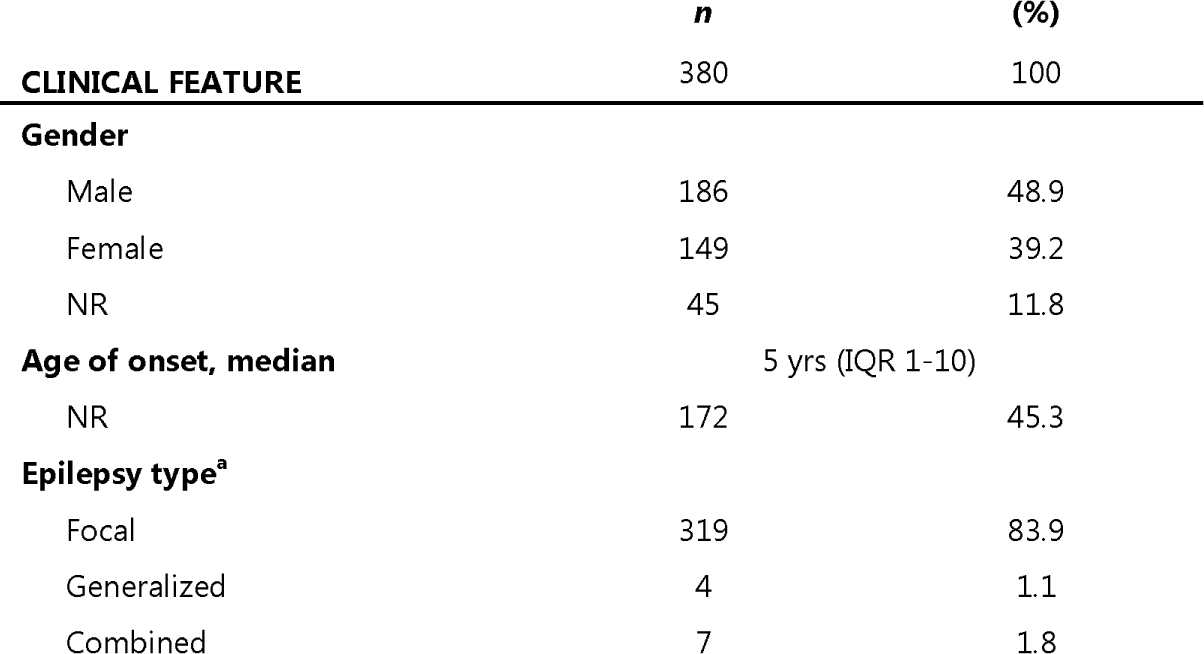
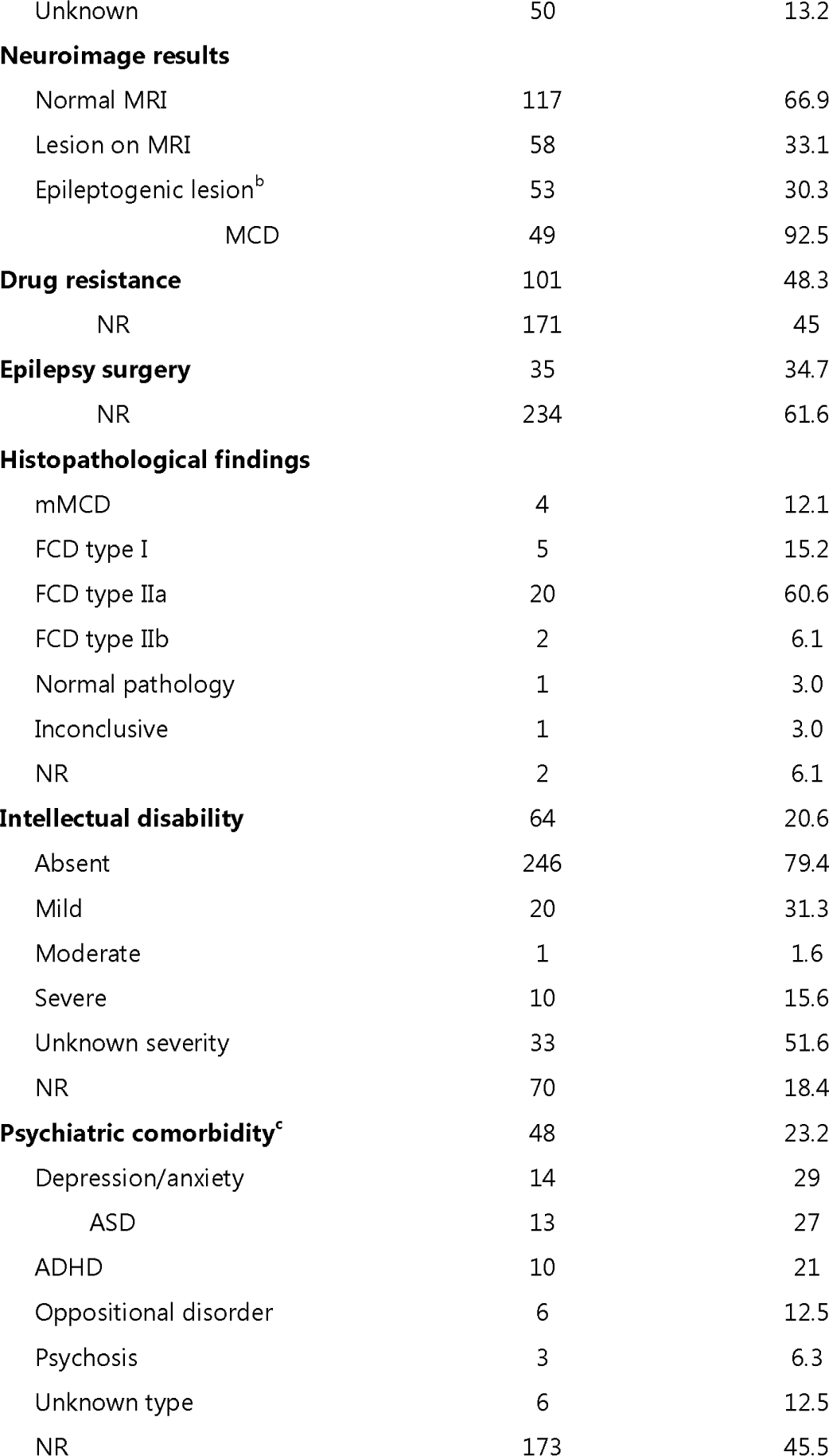
^a^Epilepsy types were classified according to the ILAE Classification of the Epilepsies 2017.^22^ ^b^Lesion causative of epilepsy, excluding nonspecific lesions (diffuse atrophy and nonspecific deep subcortical white matter disease). ^c^The number of individuals and percentages are greater than the total count since, in four individuals, two psychiatric comorbidities co-occurred. ASD: autism spectrum disorder; FCD; focal cortical dysplasia; MCD: malformation of cortical development; NR: not reported.

|  | *n* | (%) |
| --- | --- | --- |
| **CLINICAL FEATURE** | 380 | 100 |
| **Gender** |  |  |
| Male | 186 | 48.9 |
| Female | 149 | 39.2 |
| NR | 45 | 11.8 |
| **Age of onset, median** | 5 yrs (IQR 1-10) |  |
| NR | 172 | 45.3 |
| **Epilepsy type<sup>a</sup>** |  |  |
| Focal | 319 | 83.9 |
| Generalized | 4 | 1.1 |
| Combined | 7 | 1.8 |
| Unknown | 50 | 13.2 |
| **Neuroimage results** |  |  |
| Normal MRI | 117 | 66.9 |
| Lesion on MRI | 58 | 33.1 |
| Epileptogenic lesion<sup>b</sup> | 53 | 30.3 |
| MCD | 49 | 92.5 |
| **Drug resistance** | 101 | 48.3 |
| NR | 171 | 45 |
| **Epilepsy surgery** | 35 | 34.7 |
| NR | 234 | 61.6 |
| **Histopathological findings** |  |  |
| mMCD | 4 | 12.1 |
| FCD type I | 5 | 15.2 |
| FCD type IIa | 20 | 60.6 |
| FCD type IIb | 2 | 6.1 |
| Normal pathology | 1 | 3.0 |
| Inconclusive | 1 | 3.0 |
| NR | 2 | 6.1 |
| **Intellectual disability** | 64 | 20.6 |
| Absent | 246 | 79.4 |
| Mild | 20 | 31.3 |
| Moderate | 1 | 1.6 |
| Severe | 10 | 15.6 |
| Unknown severity | 33 | 51.6 |
| NR | 70 | 18.4 |
| **Psychiatric comorbidity<sup>c</sup>** | 48 | 23.2 |
| Depression/anxiety | 14 | 29 |
| ASD | 13 | 27 |
| ADHD | 10 | 21 |
| Oppositional disorder | 6 | 12.5 |
| Psychosis | 3 | 6.3 |
| Unknown type | 6 | 12.5 |
| NR | 173 | 45.5 |

Across all families, intellectual disability was reported exclusively in individuals with epilepsy and a variant in *DEPDC5*, with a frequency of 20.65% (*n=*64/310) (Figure 2C). Psychiatric comorbidities were identified in 23% (*n=*48/207) of individuals with *DEPDC5*-related epilepsy, with depression/anxiety and autism spectrum disorder being the most common (Table 1). Only one unaffected variant carrier had autism spectrum disorder (1/89 with reported psychiatric status). Non-carrier family members with epilepsy did not exhibit psychiatric comorbidity (11/14 with reported presence or absence of psychiatric phenotype).

### Associations among phenotypes

Individuals with drug-resistant epilepsy had an 8.3-fold enrichment of intellectual disability (ID) compared to individuals without drug-resistant epilepsy (Figure 3A). In addition, individuals with drug-resistant epilepsy had a 3.7-fold enrichment of lesions on MRI compared to drug responders (Figure 3A). Drug resistance (DR), intellectual disability, and the presence of a lesion on MRI were more prevalent in individuals with an early seizure onset (Figure 3B-C, E). We did not observe a statistically significant difference in the ratio of protein truncating variants (PTVs) variants compared to “other” variants such as INDELs or missense variants (see methods for details) between individuals with early seizure onset (≤1 year) and those with late seizure onset (≥10 years) (Early onset_ptv_=47, Earlyonset_otherVariants_ =10, Lateonset_ptv_=56, Lateonset_otherVariants_= 5; Fisher’s Exact Test: OR=2.4, 95%CI [0.68-9.47], *P* = 0.169). No significant association was identified between psychiatric comorbidities (Figure 3D) and seizure onset. Individuals who underwent epilepsy surgery had an earlier age of seizure onset (Figure 3F).

**Figure 3:**
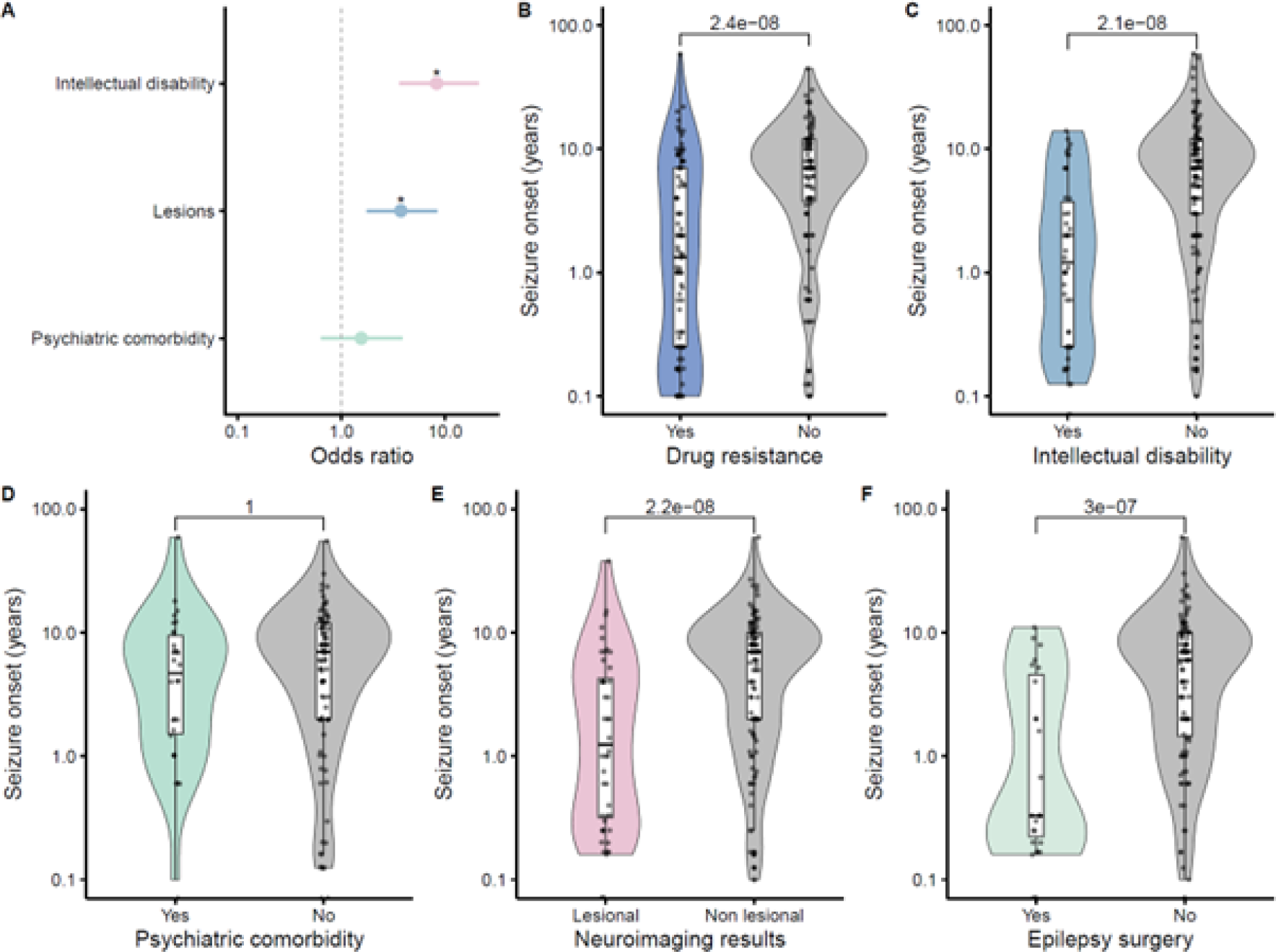
Clinical feature associations in *DEPDC5*-associated epilepsy. A) Forest plot indicating a significant association of drug resistance with the presence of lesions (Fisher’s Exact Test 95%CI [1.8-8.3]), *P* = 7.4 x 10□□), and intellectual disability (Fisher’s Exact Test 95%CI [3.6-21.0], *P*= 2.4 x 10^-8^) in variant carriers with epilepsy. B) Violin plot contrasting seizure onset ages between drug-resistant and non-drug-resistant individuals, revealing an earlier onset in the former group. Drug resistance (DR): Wilcoxon Rank sum test, *P* = 2.4e-08, *W*=2220.5, DR group size= 97, no-DR group size=91, DR_Median_=1.33, DR_IQR_=6.75, No-DR_Median_=7, No-DR_IQR_=8.2. C-F) As in B, violin plots highlight the trend of earlier seizure onset associated with intellectual disability, lesions on MRI, and epilepsy surgery, respectively. Intellectual disability (ID): Wilcoxon Rank sum test, *P* = 2.1e-08, *W*=1525.5, ID group size=54, No-ID group size=127, ID_Median_=0.9, ID_IQR_=2.7, No-ID_Median_=7, No-ID_IQR_=9.5. Lesion on MRI: Wilcoxon Rank sum test, *P* =2.2e-08, *W*=4914.5, Lesion group size=58, no Lesion group size = 109, Lesion_Median_ =0.5, Lesion_IQR_=3.8, No-Lesion_Median_=7, No-Lesion_IQR_=8. Psychiatric comorbidity (Psy): Wilcoxon Rank sum test, *P*=1, *W*= 1192.5, Psy group size=31, no Psy group size=89, Psy_Median_ = 4, Psy_IQR_ = 7.8, No-Psy_Median_=7, No-Psy_IQR_=9. Surgery (Surg): Wilcoxon Rank sum test, *P*=3e-07, *W*=657.5, Surg group size=34, no Surg group size=103, Surg _Median_=0.2, Surg _IQR_=1.9, No-Surg_Median_=6, No-Surg _IQR_=8.8

### SUDEP frequency

Studies after 2015 have introduced SUDEP as an outcome after the first report of autopsy-confirmed SUDEP cases in a family with *DEPDC5*-related epilepsy.^23^ All families included in our review had information on mortality except for one. From the overall cohort, 84% of all deceased individuals (*n*= 63/75) had epilepsy, 39.68% (*n=*25/63) of whom had confirmed or inferred *DEPDC5* variants, one individual was non-carrier, and 60.31% (*n=*37/63) had an unknown genotype. SUDEP was reported as the cause of death in 6.5% (*n=*11/169) families with reported mortality and in 16% (*n=*4/25) of deceased individuals with *DEPDC5*-related epilepsy (confirmed or inferred carriers). These four individuals had a median age of death of 50 years (IQR 43.25-52). Among all deceased individuals with epilepsy, regardless of genotype, SUDEP accounted for 20.6% (*n=*13/63) of deaths, with a median age of death of 43 years (IQR 25.5-53.75). This group included individuals without confirmed *DEPDC5* variants and those without genotypification. However, the cause of death was unknown for 76.2% (*n=*48/63) of deceased individuals with reported epilepsy. Response to antiseizure medications was only reported for 3/13 individuals with reported SUDEP, all being *DEPDC5* variant carriers.

## Discussion

### Clinical spectrum of a diverse cohort of familial *DEPDC5*-related epilepsy

We present the most comprehensive literature review to date on *DEPDC5*-related epilepsies, encompassing 170 families and 586 individuals carrying *DEPDC5* heterozygous variants. This cohort, six times larger than prior studies, includes individuals from diverse genetic backgrounds, offering a robust dataset for analysis. We used this data to identify novel genotype-phenotype associations and updated epilepsy penetrance estimates. Notably, earlier seizure onset strongly correlates with disease severity, as evidenced by significant associations with drug resistance, lesional MRI findings, and intellectual disability.

Our estimated penetrance of 64.9% aligns with prior smaller cohort estimates, although slight variations in study methodology, such as the inclusion of individuals with isolated autism spectrum disorder without epilepsy in earlier calculations, may account for differences.^5,15^ Notably, we excluded isolated non-epilepsy phenotypes due to inconsistent reporting across studies. The stable penetrance across multiplex and non-multiplex families suggests that the variants alone confer consistent disease risk independent of familial clustering. To our knowledge, this is the first study to examine epilepsy penetrance in multiplex families with *DEPDC5* variants. Clinically, the cumulative incidence showed a marked increase in the first decade of life, with 76% of affected individuals experiencing seizure onset by age 10. Accurate penetrance estimates will require future studies to report the age of unaffected carriers, a limitation in existing data where only three such cases included age at assessment.

Intellectual disability and psychiatric comorbidities were not reported in non-variant carrier individuals with epilepsy, suggesting a potential intrinsic risk conferred by *DEPDC5* variants. The prevalence of intellectual disability and psychiatric disorders in *DEPDC5* carriers without epilepsy remains unknown, highlighting the need for comprehensive phenotyping to inform genetic counselling and family planning, particularly as most *DEPDC5* variants are inherited.

Our inclusion of individuals of Asian ancestry improves the generalizability of findings compared to prior studies focused predominantly on European populations.^15,24–26^ However, data from Latin American, African, and other underrepresented groups remains absent. Future studies should prioritise these groups to refine our understanding of ancestry-specific effects on phenotype severity and penetrance.

A 26-percentage-point increase in the frequency of structural lesions detected on MRI, primarily malformations of cortical development (MCD), was noted when compared to prior pooled analyses, likely reflecting advancements in imaging techniques and post-processing methods.^13,27^ Recent studies demonstrate the importance of repeat imaging and advanced modalities, such as ^18^F-FDG PET, for detecting subtle pathologies like bottom-of-sulcus dysplasia, which were initially missed in 68% of cases but later diagnosed.^28^. These findings underscore the evolving role of imaging in refining diagnosis and guiding management of *DEPDC5*-related epilepsy.

### Drug resistance and epilepsy surgery

Drug resistance occurred in 48% of individuals with *DEPDC5* variants, higher than the 19% reported in population-based paediatric epilepsy studies and the 36% in clinic-based cohorts.^29,30^ This aligns with other Mendelian forms of epilepsy, often associated with refractory disease.^31^ Among individuals with *DEPDC5* variants, 84% had focal epilepsy, comparable to a meta-analysis reporting a 35.6% prevalence of drug resistance in focal epilepsy (95%CI: [29.3–42.2]).^32^ However, this high rate may reflect ascertainment bias, as most probands were identified at centres specialising in refractory epilepsy. Treatment response data were unavailable for 45% of individuals, emphasising the need for prospective, community-based studies to provide a more balanced understanding.

Epilepsy surgery was performed in 35% of drug-resistant individuals, a promising proportion given the well-documented underutilisation of surgical interventions in such cases.^33^ Despite MRI revealing malformations of cortical development (MCD) in 43.9% (*n=*40/91) of those with drug resistance, only a subset underwent surgery, suggesting gaps in referral practices. Despite favourable surgical outcomes in most cases (nearly 90%), the underutilisation of surgery suggests a need for improved referral practices and clinician awareness. Histopathology confirmed focal cortical dysplasia (FCD) in 77% of cases, consistent with reported high success rates for FCD resection.^32^ A recent systematic review of 44 patients with *DEPDC5* variants undergoing focal resections reported seizure freedom in 78%, supporting our findings.^34^ However, reporting bias and focusing on successful cases may influence these outcomes. Addressing barriers to surgical intervention, such as limited access to advanced imaging and specialised care centres, could significantly improve outcomes in *DEPDC5*-related epilepsy.

### Younger age of onset as a marker of disease severity

Earlier onset of *DEPDC5*-related epilepsy was associated with intellectual disability, lesional MRI, drug resistance, and epilepsy surgery. These findings align with prior studies in diverse paediatric epilepsy populations. A meta-analysis of 17 studies identified younger onset as a strong predictor of drug resistance,^30^ while a prospective study found drug resistance correlated with lower cognitive performance in early-onset cases.^35^ However, a retrospective study of epilepsy onset in the first year reported outcomes were more influenced by aetiology than age of onset.^36^ With 25% of our cohort experiencing epilepsy in the first year, our findings suggest that early seizure onset in familial *DEPDC5*-related epilepsy is a marker of severity. This underscores the need for prospective studies to refine this population’s risk stratification and outcome measures. Prompt evaluation for epilepsy surgery may benefit children with *DEPDC5*-related epilepsy, even before drug resistance develops. A retrospective study of 882 children with drug-resistant epilepsy reported postoperative improvements in cognitive function supporting early surgical intervention.^37^The positive outcomes after epilepsy surgery observed in our review underscore the critical role of early evaluation for epilepsy surgery in individuals with *DEPDC5* variants, particularly for children with early-onset epilepsy, especially because surgery may offer cognitive benefits in the long term.^37^

### SUDEP risk

*DEPDC5* variants are associated with increased SUDEP risk.^17^ An exome-based analysis of 57 SUDEP cases identified five cases carrying a *DEPDC5* variant qualifying as SUDEP risk-conferring when compared to 2,936 controls.^38^ Furthermore, *DEPDC5* gene ranked first as “probably damaging” among 72 epilepsy-related genes analysed, highlighting its association with SUDEP.^38^ In our review, SUDEP occurred in 6.5% of families, of which four individuals had confirmed or inferred *DEPDC5*-variants. The median age of SUDEP was 50 years. Baldassari et al. reported SUDEP in 9% of families from an international cohort.^13^ Inconsistent reports on follow-up time hinder reliable estimates of SUDEP incidence. Mechanisms linking *DEPDC5* to SUDEP are unclear.^39,40^ In a mouse model, the deletion of *DEPDC5* in excitatory neurons caused baseline respiratory dysfunction and frequent spontaneous tonic-clonic seizures with peri-ictal apnoea, leading to SUDEP in almost all of these *DEPDC5* knockout mice.^40^ A clinical report exploring ictal apnoea in 29 individuals with focal epilepsy found pathogenic *DEPDC5* variants in 5/14 patients with ictal central apnoea—a potential SUDEP marker—^41,42^ versus 0/15 without apnea.^43^ All seizures in *DEPDC5*-related epilepsy patients exhibited ictal central apnea.^43^ Whether *DEPDC5* variants increase SUDEP risk via respiratory dysfunction or SUDEP results from drug resistance and high frequency of nocturnal tonic-clonic seizures remains unresolved. The strong association with SUDEP risk, supported by genomic studies and animal models, highlights the importance of genetic testing and targeted interventions in affected families. Mechanistic insights into *DEPDC5*’s role in ictal apnoea and respiratory dysfunction, possibly through downstream effects of mTOR pathway dysregulation, may open new risk stratification and prevention avenues. Prospective, deep-phenotypification studies with sufficient follow-up are needed.

### Limitations

This retrospective review is inherently limited to available genotype and phenotype information from families at the time of publication. Not all family members may have been offered or willing to participate in genetic testing, and some individuals may develop epilepsy after the initial report. Limited family testing and a bias towards testing individuals with known phenotypes may lead to an overestimated penetrance. Nevertheless, this study represents the most comprehensive and diverse dataset to date, making it a valuable resource for understanding familial *DEPDC5*-related epilepsy across varied ancestries, addressing the limitations of previous smaller cohorts.

Publication bias likely influenced the favourable outcomes observed with epilepsy surgery, as studies with less successful results may be underreported. Our analysis of clinical feature associations is limited by the lack of adjustment for disease phenotypes and potential confounding factors, and the associations observed may be influenced by a bias toward more severe, earlier-presenting cases. Additionally, SUDEP observations were not included in the clinical analysis, as most cases occurred in individuals for whom genetic testing was unavailable. While we screened additional literature to identify further reports, it is possible that SUDEP occurred after the data was published.

### Future directions

Prospective, genetically diverse cohorts of *DEPDC5* variant carrier families that identify predictors of severity, outcomes, and SUDEP will help inform effective counselling approaches and precision medicine trials. Developing risk stratification tools to identify individuals at higher risk for drug resistance and SUDEP will allow more focused preventive strategies and enhanced patient care. Underpinning the frequency and phenotype of comorbidities and non-CNS manifestations of these variants will provide a comprehensive assessment of individuals and families harbouring *DEPDC5* variants, offering quality and precision-medicine comprehensive care for individuals with these variants.

## Conclusion

This literature update not only corroborates previous penetrance estimates but also identifies early seizure onset as a critical prognostic marker for disease severity in *DEPDC5*-related epilepsy. Our findings, derived from the largest dataset to date, including recent data from Asian populations, offer valuable insights into this condition’s clinical management and prognosis.

## Supplementary material

Supplementary material is available at *Brain* online.

## Data Availability

All data produced in the present work are contained in the manuscript or Supplementary material

## References

1. Saxton RA, Sabatini DM. mTOR Signaling in Growth, Metabolism, and Disease. Cell. 2017;168(6):960–976. doi:10.1016/j.cell.2017.02.004

2. Iffland PH, Carson V, Bordey A, Crino PB. GATORopathies: The role of amino acid regulatory gene mutations in epilepsy and cortical malformations. Epilepsia. 2019;60(11):2163–2173. doi:10.1111/epi.16370

3. Lipton JO, Sahin M. The Neurology of mTOR. Neuron. 2014;84(2):275–291. doi:10.1016/j.neuron.2014.09.034

4. Ishida S, Picard F, Rudolf G, et al. Mutations of DEPDC5 cause autosomal dominant focal epilepsies. Nat Genet. 2013;45(5):552–555. doi:10.1038/ng.2601

5. Dibbens LM, De Vries B, Donatello S, et al. Mutations in DEPDC5 cause familial focal epilepsy with variable foci. Nat Genet. 2013;45(5):546–551. doi:10.1038/ng.2599

6. Picard F, Makrythanasis P, Navarro V, et al. DEPDC5 mutations in families presenting as autosomal dominant nocturnal frontal lobe epilepsy. Neurology. 2014;82(23):2101–2106. doi:10.1212/WNL.0000000000000488

7. Carvill GL, Crompton DE, Regan BM, et al. Epileptic spasms are a feature of DEPDC5 mTORopathy. Neurol Genet. 2015;1(2). doi:10.1212/NXG.0000000000000016

8. Lal D, Reinthaler EM, Schubert J, et al. DEPDC5 mutations in genetic focal epilepsies of childhood. Ann Neurol. 2014;75(5):788–792. doi:10.1002/ana.24127

9. Baulac S, Ishida S, Marsan E, et al. Familial focal epilepsy with focal cortical dysplasia due to DEPDC 5 mutations. Ann Neurol. 2015;77(4):675–683. doi:10.1002/ana.24368

10. Scheffer IE, Heron SE, Regan BM, et al. Mutations in mammalian target of rapamycin regulator DEPDC5 cause focal epilepsy with brain malformations. Ann Neurol. 2014;75(5):782–787. doi:10.1002/ana.24126

11. Baldassari S, Ribierre T, Marsan E, et al. Dissecting the genetic basis of focal cortical dysplasia: a large cohort study. Acta Neuropathol. 2019;138(6):885–900. doi:10.1007/s00401-019-02061-5

12. D’Gama AM, Geng Y, Couto JA, et al. Mammalian target of rapamycin pathway mutations cause hemimegalencephaly and focal cortical dysplasia. Ann Neurol. 2015;77(4):720–725. doi:10.1002/ana.24357

13. Baldassari S, Picard F, Verbeek NE, et al. The landscape of epilepsy-related GATOR1 variants. Genetics in Medicine. 2019;21(2):398–408. doi:10.1038/s41436-018-0060-2

14. Symonds JD, Zuberi SM, Stewart K, et al. Incidence and phenotypes of childhood-onset genetic epilepsies: A prospective population-based national cohort. Brain. 2019;142(8):2303–2318. doi:10.1093/brain/awz195

15. Gu C, Wei X, Yan D, et al. DEPDC5 plays a vital role in epilepsy: Genotypic and phenotypic features in cohort and literature. Epileptic Disorders. 2024;26(3):341–349. doi:10.1002/epd2.20223

16. Hadzsiev K, Hegyi M, Fogarasi A, et al. Observation of a Possible Successful Treatment of DEPDC5 -Related Epilepsy with mTOR Inhibitor. Neuropediatrics. 2022;54(5):344–346. doi:10.1055/a-2104-1614

17. Klofas LK, Short BP, Zhou C, Carson RP. Prevention of premature death and seizures in a Depdc5 mouse epilepsy model through inhibition of mTORC1. Hum Mol Genet. 2020;29(8):1365–1377. doi:10.1093/hmg/ddaa068

18. Moloney PB, Kearney H, Benson KA, et al. Everolimus precision therapy for the GATOR1-related epilepsies: A case series. Eur J Neurol. 2023;30(10):3341–3346. doi:10.1111/ene.15975

19. Tricco AC, Lillie E, Zarin W, et al. PRISMA Extension for Scoping Reviews (PRISMA-ScR): Checklist and Explanation. Ann Intern Med. 2018;169(7):467–473. doi:10.7326/M18-0850

20. Sanders S. Multiplex-Simplex Comparisons. In: Volkmar FR, ed. Encyclopedia of Autism Spectrum Disorders. Springer New York; 2013:1960. doi:10.1007/978-1-4419-1698-3_1333

21. Team CR. R: A Language and Environment for Statistical Computing. [software] [Internet]. Vienna, Austria: R Foundation for Statistical Computing; 2017. Available from https://www.r[project.org/. R Foundation for Statistical Computing. 2022. https://www.r-project.org/

22. Scheffer IE, Berkovic S, Capovilla G, et al. ILAE classification of the epilepsies: Position paper of the ILAE Commission for Classification and Terminology. Epilepsia. 2017;58(4):512–521. doi:10.1111/epi.13709

23. Nascimento FA, Borlot F, Cossette P, Minassian BA, Andrade DM. Two Definite Cases of Sudden Unexpected Death in Epilepsy in a Family with a DEPDC5 Mutation. Neurol Genet. 2015;1(4):1–3. doi:10.1212/NXG.0000000000000028

24. Yin K, Lei X, Yan Z, et al. Clinical and genetic features of GATOR1 complex-associated epilepsy. Published online 2023:784–790. doi:10.1136/jmedgenet-2021-108364

25. Wang H, Liu W, Zhang Y, Liu Q, Cai L, Jiang Y. Seizure features and outcomes in 50 children with GATOR1 variants: A retrospective study, more favorable for epilepsy surgery. Epilepsia Open. 2023;8(3):969–979. doi:10.1002/epi4.12770

26. Wang Y, Niu W, Shi H, et al. A novel variation in DEPDC5 causing familial focal epilepsy with variable foci. Front Genet. 2024;15. doi:10.3389/fgene.2024.1414259

27. Kanber B, Vos SB, de Tisi J, et al. Detection of covert lesions in focal epilepsy using computational analysis of multimodal magnetic resonance imaging data. Epilepsia. 2021;62(3):807–816. doi:10.1111/epi.16836

28. Macdonald-Laurs E, Warren AEL, Francis P, et al. The clinical, imaging, pathological and genetic landscape of bottom-of-sulcus dysplasia. Brain. 2024;147(4):1264–1277. doi:10.1093/brain/awad379

29. Sillanpää M, Schmidt D. Natural history of treated childhood-onset epilepsy: prospective, long-term population-based study. Brain. 2006;129(Pt 3):617–624. doi:10.1093/brain/awh726

30. Sultana B, Panzini MA, Veilleux Carpentier A, et al. Incidence and Prevalence of Drug-Resistant Epilepsy. Neurology. 2021;96(17):805–817. doi:10.1212/WNL.0000000000011839

31. Ayoub D, Jaafar F, Al-Hajje A, et al. Predictors of drug-resistant epilepsy in childhood epilepsy syndromes: A subgroup analysis from a prospective cohort study. Epilepsia. 2024;65(10):2995–3009. doi:10.1111/epi.18100

32. Willard A, Antonic-Baker A, Chen Z, O’Brien TJ, Kwan P, Perucca P. Seizure Outcome After Surgery for MRI-Diagnosed Focal Cortical Dysplasia: A Systematic Review and Meta-analysis. Neurology. 2022;98(3):e236–e248. doi:10.1212/WNL.0000000000013066

33. Solli E, Colwell NA, Say I, et al. Deciphering the surgical treatment gap for drug[resistant epilepsy (DRE): A literature review. Epilepsia. 2020;61(7):1352–1364. doi:10.1111/epi.16572

34. McGinley C, Teti S, Hofmann K, et al. Seizure Control Outcomes following Resection of Cortical Dysplasia in Patients with DEPDC5 Variants: A Systematic Review and Individual Patient Data Analysis. Neuropediatrics. 2024;55(1):1–8. doi:10.1055/a-2213-8584

35. Berg AT, Zelko FA, Levy SR, Testa FM. Age at onset of epilepsy, pharmacoresistance, and cognitive outcomes: A prospective cohort study. Neurology. 2012;79(13):1384–1391. doi:10.1212/WNL.0b013e31826c1b55

36. Gaily E, Lommi M, Lapatto R, Lehesjoki AE. Incidence and outcome of epilepsy syndromes with onset in the first year of life: A retrospective population-based study. Epilepsia. 2016;57(10):1594–1601. doi:10.1111/epi.13514

37. Eriksson MH, Prentice F, Piper RJ, et al. Long-term neuropsychological trajectories in children with epilepsy: does surgery halt decline? Brain. 2024;147(8):2791–2802. doi:10.1093/brain/awae121

38. Bagnall RD, Crompton DE, Petrovski S, et al. Exome-based analysis of cardiac arrhythmia, respiratory control, and epilepsy genes in sudden unexpected death in epilepsy. Ann Neurol. 2016;79(4):522–534. doi:10.1002/ana.24596

39. Bacq A, Roussel D, Bonduelle T, et al. Cardiac Investigations in Sudden Unexpected Death in DEPDC5-Related Epilepsy. Ann Neurol. 2022;91(1):101–116. doi:10.1002/ana.26256

40. Kao HY, Yao Y, Yang T, et al. Sudden Unexpected Death in Epilepsy and Respiratory Defects in a Mouse Model of DEPDC5-Related Epilepsy. Ann Neurol. 2023;94(5):812–824. doi:10.1002/ana.26773

41. Vilella L, Lacuey N, Hampson JP, et al. Postconvulsive central apnea as a biomarker for sudden unexpected death in epilepsy (SUDEP). Neurology. 2019;92(3):E171–E182. doi:10.1212/WNL.0000000000006785

42. Lacuey N, Zonjy B, Hampson JP, et al. The incidence and significance of periictal apnea in epileptic seizures. Epilepsia. 2018;59(3):573–582. doi:10.1111/epi.14006

43. Meletti S, Duma GM, Burani M, et al. Ictal and Postictal Central Apnea in DEPDC5 - Related Epilepsy. Neurol Genet. 2024;10(5). doi:10.1212/NXG.0000000000200183

