## Supplementary Material for "Insights into *DEPDC5*-Related Epilepsy from 586 people: Variant Penetrance, Phenotypic Spectrum, and Treatment Outcomes"

### Supplementary Methods

#### Publication search strategy - Extended

A scoping literature review was conducted following the PRISMA-ScR (Preferred Reporting Items for Systematic Reviews and Meta-Analyses Extension for Scoping Reviews)[1] checklist (Table 1), considering all studies listed on PubMed until August 31, 2024. We used the gene-specific term "*DEPDC5*" as the keyword for the PubMed search. The publications were reviewed independently in two steps by two investigators (EAB and MOU) as follows: (a) manual title and abstract screening to remove non-English studies, duplicated studies (including commentary or response letters), reviews, animal studies, cellular level studies, studies reporting on genes other than *DEPDC5*, and studies focused on non-neurological phenotypes; (b) manual full-text review to select only studies where genotype and phenotype information was available for families, such that families had ≥2 individuals with a heterozygous germline variant in the gene *DEPDC5* and at least one individual with a heterozygous *DEPDC5* variant with an epilepsy phenotype. We excluded studies that reported individual patients without family information or sporadic cases (i.e., due to de novo or somatic variants) and studies reporting disease association with a recessive mode of inheritance, as this was only reported once and described a distinct disorder spectrum.[2] We eliminated duplicate families used in multiple publications by reviewing citations to previous studies, authors/institutions, genetic information, pedigree structure, and phenotype information. The relevant genotypic and phenotypic data was extracted independently by the two reviewers (EAB and MOU) in an Excel form developed with the senior author (DL) and two senior epileptologists (SL, IN). The overall publication screening process is detailed in Figure 1.

#### Data collection and organization extended

We extracted genotype and phenotype information from the final 33 publications identified in the literature review (the full list is given in Supplementary Table 2). Variables collected included the number of individuals carrying heterozygous variants in *DEPDC5* with and without a documented epilepsy phenotype and information on the genetic variant type (missense variant or protein-truncating variant, PTV) utilising cDNA change and amino acid change. Obligate carriers, either denoted by the original studies or inferred through pedigree analysis (following Otto and Horimoto, 2012)[3] by a co-author with high expertise in genetic variation models (CL). Genotype data was collected as reported by the authors utilizing cDNA change and amino acid change. All identified genetic variants were categorized into three groups based on their molecular characteristics. Variants leading to a premature stop codon, frameshift mutations resul ting in a premature stop codon, splice-site variants and single and multiple whole-exon deletions were classified as protein-truncating variants (PTVs). Single-nucleotide changes in the DNA sequence that result in the substitution of one amino acid for another were grouped together with in-frame insertions and deletions (INDELs) and are referred to as "Other" variants.

We meticulously extracted clinical information for all the individuals with documented epilepsy, fulfilling the International League Against Epilepsy (ILAE) definition of epilepsy[4] regardless of their variant status. Clinical variables collected were: age of seizure onset; epilepsy type according to the latest ILAE classification[5]; drug resistance, as defined by the authors of each report; neuroimaging results reported; author reported presence of intellectual disability and/or psychiatric comorbidity, reported epilepsy surgery, histopathological report and reported classification of focal cortical dysplasia when pathology was available, sudden unexpected death in epilepsy (SUDEP) and type of SUDEP as reported by the authors, and age of death. The study adhered to the ILAE definition of epilepsy[4] and ILAE epilepsy classification[5] to ensure clinical consistency in identifying epilepsy phenotypes among *DEPDC5* variant carriers. Psychiatric comorbidity included schizophrenia, depression and anxiety, autism spectrum disorder, and attention-deficit/hyperactivity disorder (ADHD). If the family was reported in additional literature reports, we screened these reports and added relevant clinical information whenever available (Supplementary Table 2). To determine SUDEP frequency, we included information from confirmed or inferred *DEPDC5* variant carriers and from family members with epilepsy and an unknown genotype. The decision to include individuals without known genotypes was based on the first report of two definite SUDEP cases without available DNA in a family with *DEPDC5*-related epilepsy.[6]The authors hypothesised that these individuals were unlikely to carry variants in other genes previously associated with SUDEP and suggested a strong likelihood of *DEPDC5* involvement in families with multiple SUDEP cases.[6]

A sub-cohort of multiplex families (*n*= 68) was created, where at least two individuals had a pathogenic *DEPDC5* variant confirmed by genetic sequencing and an epilepsy phenotype, to better understand the inheritance patterns from large families with multiple affected and/or tested offspring or across generations. For this sub-cohort, we excluded families where only the index case and parents were sequenced.

##### Statistical analysis and penetrance calculation extended

All statistical analyses were carried out in R, version 4.4.0.[7] Clinical data analysis was performed for individuals with epilepsy and confirmed or inferred variants (obligate carriers). To assess the enrichment of clinical features among patients with drug-resistant epilepsy, we employed the Fisher's Exact Test. Associations between the age of seizure onset and clinical variables collected were quantified using the Wilcoxon Rank Sum Test. The cumulative incidence of age of seizure onset was calculated by determining the proportion of individuals who had experienced their first seizure at each age. Binomial confidence intervals (95%) were computed using the standard error of the proportions, with lower and upper bounds derived from the formula:

$$p \pm 1.96 \times \sqrt{\frac{p(1-p)}{n}}$$

where $p$ is the observed proportion and n is the number of individuals with data available. These estimates were derived exclusively from the group of individuals with DEPDC5-related epilepsy, excluding variant carriers without epilepsy.

Penetrance was calculated as the total number of individuals across all families with heterozygous variants in *DEPDC5* and an epilepsy phenotype, divided by the total number of individuals across all families with heterozygous variants in *DEPDC5* (with or without an epilepsy phenotype). Penetrance calculations included confirmed and obligate carriers. The estimated penetrance for multiplex families was also calculated and compared with the penetrance across all families by two-tailed t-test.

REFERENCES:

1. Tricco AC, Lillie E, Zarin W, et al. PRISMA Extension for Scoping Reviews (PRISMA-ScR): Checklist and Explanation. *Ann Intern Med*. 2018;169(7):467-473. doi:10.7326/M18-0850
2. Ververi A, Zagaglia S, Menzies L, et al. Germline homozygous missense DEPDC5 variants cause severe refractory early-onset epilepsy, macrocephaly and bilateral polymicrogyria. *Hum Mol Genet*. 2023;32(4):580-594. doi:10.1093/hmg/ddac225
3. Otto PA, Horimoto ARVR. Penetrance rate estimation in autosomal dominant conditions. *Genet Mol Biol*. 2012;35(3):583-588. doi:10.1590/S1415-47572012005000051
4. Fisher RS, Acevedo C, Arzimanoglou A, et al. ILAE Official Report: A practical clinical definition of epilepsy. *Epilepsia*. 2014;55(4):475-482. doi:10.1111/epi.12550
5. Scheffer IE, Berkovic S, Capovilla G, et al. ILAE classification of the epilepsies: Position paper of the ILAE Commission for Classification and Terminology. *Epilepsia*. 2017;58(4):512-521. doi:10.1111/epi.13709
6. Nascimento FA, Borlot F, Cossette P, Minassian BA, Andrade DM. Two Definite Cases of Sudden Unexpected Death in Epilepsy in a Family with a DEPDC5 Mutation. *Neurol Genet*. 2015;1(4):1-3. doi:10.1212/NXG.0000000000000028
7. Team CR. R: A Language and Environment for Statistical Computing. [software] [Internet]. Vienna, Austria: R Foundation for Statistical Computing; 2017. Available from https://www.r-project.org/. R Foundation for Statistical Computing. 2022. https://www.r-project.org/

#### Supplementary Table 1. Preferred Reporting Items for Systematic reviews and Meta-Analyses extension for Scoping Reviews (PRISMA-ScR) Checklist

| SECTION | ITEM | PRISMA-ScR CHECKLIST ITEM | REPORTED ON PAGE # |
|---|---|---|---|
| **TITLE** | | | |
| **Title** | 1 | Identify the report as a scoping review. | 1 |
| **ABSTRACT** | | | |
| **Structured summary** | 2 | Provide a structured summary that includes (as applicable): background, objectives, eligibility criteria, sources of evidence, charting methods, results, and conclusions that relate to the review questions and objectives. | 1-2 |
| **INTRODUCTION** | | | |

| | | | |
|---|---|---|---|
| **Rationale** | 3 | Describe the rationale for the review in the context of what is already known. Explain why the review questions/objectives lend themselves to a scoping review approach. | 4 |
| **Objectives** | 4 | Provide an explicit statement of the questions and objectives being addressed with reference to their key elements (e.g., population or participants, concepts, and context) or other relevant key elements used to conceptualize the review questions and/or objectives. | 4 |
| **METHODS** | | | |
| **Protocol and registration** | 5 | Indicate whether a review protocol exists; state if and where it can be accessed (e.g., a Web address); and if available, provide registration information, including the registration number. | Not applied |
| **Eligibility criteria** | 6 | Specify characteristics of the sources of evidence used as eligibility criteria (e.g., years considered, language, and publication status), and provide a rationale. | 5 |
| **Information sources*** | 7 | Describe all information sources in the search (e.g., databases with dates of coverage and contact with authors to identify additional sources), as well as the date the most recent search was executed. | 5 |
| **Search** | 8 | Present the full electronic search strategy for at least 1 database, including any limits used, such that it could be repeated. | 5 |
| **Selection of sources of evidence†** | 9 | State the process for selecting sources of evidence (i.e., screening and eligibility) included in the scoping review. | 5 |
| **Data charting process‡** | 10 | Describe the methods of charting data from the included sources of evidence (e.g., calibrated forms or forms that have been tested by the team before their use, and whether data charting was done independently or in duplicate) and any processes for obtaining and confirming data from investigators. | 5 |
| **Data items** | 11 | List and define all variables for which data were sought and any assumptions and simplifications made. | 5 and Suppl. |
| **Critical appraisal of individual sources of evidence§** | 12 | If done, provide a rationale for conducting a critical appraisal of included sources of evidence; describe the methods used and how this information was used in any data synthesis (if appropriate). | Not applied |
| **Synthesis of results** | 13 | Describe the methods of handling and summarizing the data that were charted. | 5 and Suppl. |
| **RESULTS** | | | |
| **Selection of sources of evidence** | 14 | Give numbers of sources of evidence screened, assessed for eligibility, and included in the review, with reasons for exclusions at each stage, ideally using a flow diagram. | 5 |
| **Characteristics of sources of evidence** | 15 | For each source of evidence, present characteristics for which data were charted and provide the citations. | Supplementary Table 2 |
| **Critical appraisal within sources of evidence** | 16 | If done, present data on critical appraisal of included sources of evidence (see item 12). | Not applied |
| **Results of individual sources of evidence** | 17 | For each included source of evidence, present the relevant data that were charted that relate to the review questions and objectives. | Supplementary Table 2 |
| **Synthesis of results** | 18 | Summarize and/or present the charting results as they relate to the review questions and objectives. | 6-8 |
| **DISCUSSION** | | | |
| **Summary of evidence** | 19 | Summarize the main results (including an overview of concepts, themes, and types of evidence available), link to the review questions and objectives, and consider the relevance to key groups. | 8-9 |
| **Limitations** | 20 | Discuss the limitations of the scoping review process. | 11 |
| **Conclusions** | 21 | Provide a general interpretation of the results with respect to the review questions and objectives, as well as potential implications and/or next steps. | 12 |
| **FUNDING** | | | |
| **Funding** | 22 | Describe sources of funding for the included sources of evidence, as well as sources of funding for the scoping review. Describe the role of the funders of the scoping review. | 13 |

*From:* Tricco AC, Lillie E, Zarin W, O'Brien KK, Colquhoun H, Levac D, et al. PRISMA Extension for Scoping Reviews (PRISMAScR): Checklist and Explanation. Ann Intern Med. 2018;169:467–473. doi: 10.7326/M18-0850.

#### Supplementary Table 2. Summary of main genotypic and phenotypic variables extracted from each study, by family.

| PMID | Family ID | Multi-plex | Epilepsy + Variant *n* | Variant carrier *n* | Epilepsy+ Variant only confirmed | Variant only confirmed carriers | Variant Type | Variant c | Variant p | Epilepsy age onset, median yrs (*n*) | DR *n/n*[a] | ID *n/n*[a] | Psychiatric comorb. *n/n*[a] | Epilepsy surgery, *n* |
|---|---|---|---|---|---|---|---|---|---|---|---|---|---|---|
| **23542697** | **A1** | Y | 9 | 13 | 8 | 11 | PTV | | p.Tyr7* | 8 (8) | 2/6 | 2/9 | 2/9 | 0/1 |
| **23542697** | **D1** | Y | 14 | 25 | 14 | 24 | PTV | | p.Arg555* | | NR | 0/14 | 2/14 | NR |
| **23542697** | **FC1** | Y | 6 | 9 | 6 | 9 | Other variant (inframe INDEL) | | p.Phe164del | | NR | 0/6 | 0/6 | NR |
| **23542697** | **FC2** | Y | 8 | 16 | 8 | 15 | Other variant (inframe INDEL) | | p.Phe164del | | NR | 0/8 | 0/8 | NR |
| **23542697** | **FC3** | Y | 21 | 30 | 21 | 26 | Other variant (inframe INDEL) | | p.Phe164del | | NR | 0/21 | 3/21 | NR |
| **23542697** | **G** | Y | 5 | 6 | 5 | 5 | PTV | | p.Trp1466* | | NR | 0/5 | 0/5 | NR |
| **23542697** | **H** | Y | 4 | 5 | 4 | 5 | PTV | c.193+1G >A | | 0.143 (2) | 0/2 | 1/4 | 1/4 | NR |
| **23542697** | **J** | Y | 3 | 4 | 3 | 4 | PTV | | p.Arg487* | | NR | 2/3 | 0/3 | NR |
| **23542697** | **K** | Y | 4 | 4 | 3 | 3 | PTV | | p.Arg843* | | NR | 0/4 | 0/4 | NR |
| **23542697** | **L** | Y | 2 | 3 | 2 | 3 | PTV | | p.Trp1466* | | NR | 0/2 | 0/2 | NR |
| **23542697** | **M** | N | 1 | 2 | 1 | 2 | PTV | | p.Arg1268* | | NR | 0/1 | 0/1 | NR |
| **23542697** | **N** | Y | 3 | 3 | 3 | 3 | Other variant (Missense) | | p.Ser1104Leu | | NR | 0/3 | 0/3 | NR |
| **23542697** | **P** | N | 2 | 2 | 2 | 2 | Other variant (Missense) | | p.Ser1104Leu | | NR | 0/2 | 0/2 | NR |
| **23542697** | **S1** | Y | 7 | 13 | 7 | 11 | PTV | | p.Trp1369* | | NR | 1/7 | 1/7 | NR |
| **23542697** | **S2** | Y | 5 | 10 | 4 | 8 | PTV | | p.Gln1536* | | NR | 0/5 | 0/5 | NR |
| **23542701** | **L** | Y | 4 | 4 | 4 | 4 | PTV | | p.Arg328* | | NR | NR | NR | NR |
| **23542701** | **N** | Y | 6 | 10 | 6 | 10 | PTV | | p.Leu374Phefs*30 | | NR | 0/1 | NR | NR |
| **23542701** | **O** | Y | 2 | 2 | 2 | 2 | Other variant (Missense) | | p.Arg485Gln | | NR | NR | NR | NR |
| **23542701** | **Q** | N | 2 | 2 | 2 | 2 | PTV | | p.Gln372* | | NR | 0/1 | NR | NR |
| **23542701** | **S** | Y | 8 | 14 | 7 | 12 | PTV | | p.Arg239* | 0.6 (3) | 3/3 | 1/3 | 2/3 | 2/3 |
| **24283814** | **1** | Y | 5 | 12 | 5 | 11 | PTV | | p.Arg843* | 12 (5) | NR | NR | 1/1 | NR |
| **24283814** | **2** | N | 1 | 2 | 1 | 2 | PTV | | p.Arg843* | 1.5 (1) | NR | NR | NR | NR |
| **24283814** | **3** | Y | 2 | 4 | 2 | 3 | PTV | | p.Arg843* | 5 (2) | NR | NR | NR | NR |
| **24585383** | **A** | Y | 6 | 10 | 6 | 10 | PTV | | p.Gln140* | 8.5 (6) | 1/6 | 1/6 | 2/6 | 0/3 |

| | | | | | | | | | | | | | | |
|---|---|---|---|---|---|---|---|---|---|---|---|---|---|---|
| **24585383** | **C** | N | 2 | 2 | 2 | 2 | PTV | c.279+1G>A | | 9.5 (2) | 0/1 | 0/2 | 0/2 | NR |
| **24591017** | **1** | N | 1 | 2 | 1 | 2 | PTV | | p.I1139Metfs*24 | 4.8 (1) | 0/1 | NR | NR | NR |
| **24591017** | **2** | Y | 2 | 4 | 2 | 4 | PTV | c.59-1G>C | | 5.75 (2) | 0/2 | NR | NR | NR |
| **24591017** | **3** | Y | 3 | 4 | 3 | 4 | PTV | | p.Arg865* | 1.625 (2) | 1/3 | 1/2 | NR | NR |
| **24591017** | **5** | N | 1 | 3 | 1 | 3 | Other variant (Missense) | | p.Val272Leu | | 0/1 | NR | NR | NR |
| **24591017** | **6** | N | 1 | 2 | 1 | 2 | Other variant (Missense) | | p.Val90Ile | | 0/1 | NR | NR | NR |
| **24591017** | **7** | N | 1 | 2 | 1 | 2 | Other variant (Missense) | | p.Ser1153Gly | | 0/1 | NR | NR | NR |
| **24814846** | **1** | Y | 2 | 2 | 2 | 2 | PTV | c.2355-2A>G | | 9.5 (2) | 1/2 | 0/2 | 0/2 | NR |
| **24814846** | **2** | Y | 2 | 3 | 2 | 3 | PTV | | p.Arg487* | 2.25 (2) | 2/2 | 2/2 | 1/2 | NR |
| **24814846** | **3** | Y | 2 | 3 | 2 | 3 | PTV | | p.Arg1087* | 15.5 (2) | 2/2 | 0/2 | 1/2 | NR |
| **24814846** | **4** | Y | 3 | 3 | 3 | 3 | PTV | | p.Trp1369* | 8 (3) | 2/2 | 1/3 | 0/3 | NR |
| **25623524** | **2** | Y | 2 | 2 | 2 | 2 | PTV | c.484-1G>A | | 1.6 (1) | 1/2 | 0/2 | 1/1 | 1/1 |
| **25623524** | **4** | Y | 2 | 2 | 2 | 2 | PTV | | p.Arg587* | 2.15 | 2/2 | 0/2 | 0/2 | 2/2 |
| **26000329** | **NA** | Y | 4 | 5 | 4 | 5 | PTV | | p.Arg555* | 12 (5) | 2/3 | 0/2 | NR | 2/2 |
| **26216793** | **NA** | N | 2 | 2 | 2 | 2 | PTV | | p.Tyr306* | 10 (2) | 0/2 | 0/2 | 0/2 | 0/2 |
| **26505888** | **11** | Y | 2 | 3 | 2 | 3 | PTV | c.3265-3C>T | | | NR | 0/2 | NR | NR |
| **26505888** | **12** | Y | 2 | 3 | 2 | 3 | PTV | | p.Val367Glyfs*40 | | NR | 0/2 | NR | NR |
| **26505888** | **13** | Y | 3 | 4 | 3 | 4 | Other variant (Missense) | | p.Gln542Pro | | NR | 0/3 | NR | NR |
| **26505888** | **14** | N | 1 | 3 | 1 | 3 | PTV | | p.Gln176* | | NR | 1/1 | NR | NR |
| **26505888** | **15** | Y | 2 | 3 | 2 | 3 | PTV | | p.Arg422* | | NR | 0/2 | NR | NR |
| **26505888** | **16** | N | 1 | 2 | 1 | 2 | PTV | c.4033+5A>G | | | NR | 0/1 | NR | NR |
| **26505888** | **17** | Y | 2 | 3 | 2 | 3 | Other variant (Missense) | | p.Ser1154Phe | | NR | 0/2 | NR | NR |
| **26505888** | **18** | N | 1 | 2 | 1 | 2 | PTV | | p.Arg165Tyrfs*14 | 10 (1) | 1/1 | 0/1 | NR | NR |
| **26505888** | **19** | N | 1 | 2 | 1 | 2 | Other variant (Missense) | | p.Thr1081Pro | | NR | 1/1 | NR | NR |
| **26505888** | **21** | Y | 2 | 3 | 2 | 3 | PTV | | p.Trp145* | | NR | 0/2 | NR | NR |
| **26505888** | **22** | N | 1 | 2 | 1 | 2 | PTV | | p.Arg1332* | | NR | 0/1 | NR | NR |
| **26505888** | **23** | Y | 2 | 2 | 2 | 2 | PTV | | p.Arg243* | 3 (2) | 1/2 | 2/2 | 0/1 | NR |
| **26505888** | **24** | N | 1 | 2 | 1 | 2 | Other variant (Missense) | | P.His214Asp | | NR | 1/1 | NR | NR |

| | | | | | | | | | | | | | | |
|---|---|---|---|---|---|---|---|---|---|---|---|---|---|---|
| **26505888** | **25** | N | 1 | 2 | 1 | 2 | Other variant (Missense) | | p.Gln54Pro | | NR | 1/1 | NR | NR |
| **26505888** | **26** | N | 1 | 2 | 1 | 2 | Other variant (Missense) | | p.Arg1268Gln | | NR | 0/1 | NR | NR |
| **26505888** | **28** | Y | 6 | 10 | 6 | 10 | PTV | | p.Thr329Leufs*7 | | NR | 0/6 | NR | NR |
| **27066544** | **B** | Y | 3 | 3 | 3 | 3 | PTV | | p.Leu1371Argfs*14 | 9 (3) | NR | 0/3 | NR | NR |
| **27066544** | **C** | N | 2 | 2 | 2 | 2 | PTV | | p.Tyr306* | 6.25 (2) | 1/1 | NR | NR | NR |
| **27066554** | **B** | N | 2 | 2 | 2 | 2 | PTV | | p.Gln519* | 0.25 (1) | 1/1 | 1/2 | 1/1 | 0/1 |
| **27066554** | **D** | Y | 3 | 3 | 3 | 3 | Other variant (Missense) | | p.Tyr281Phe | 0.125 (1) | 1/1 | 1/3 | NR | 1/1 |
| **27066565** | **NA** | Y | 8 | 9 | 6 | 7 | PTV | | p.Gln216* | 10 (8) | 2/7 | 0/8 | 1/8 | NR |
| **27173016** | **A** | Y | 4 | 4 | 4 | 4 | PTV | | p.Gln465* | | NR | NR | NR | NR |
| **27173016** | **D** | Y | 2 | 2 | 2 | 2 | PTV | | p.Leu584Phefs*12 | | NR | NR | NR | NR |
| **28170089** | **A** | Y | 5 | 5 | 5 | 5 | PTV | c.4203+2 T>A | | 13.5 (4) | 1/4 | 0/1 | NR | NR |
| **28170089** | **C** | N | 1 | 2 | 1 | 2 | Other variant (Missense) | | p.Asp967Asn | 15 (1) | 0/1 | NR | NR | NR |
| **28170089** | **E** | N | 1 | 3 | 1 | 2 | Other variant (Missense) | | p.Arg1586Trp | 2 (1) | 0/1 | NR | NR | NR |
| **28199897** | **8** | Y | 2 | 3 | 2 | 3 | PTV | | p.Gln140* | 2 (1) | 0/1 | 0/1 | NR | NR |
| **29708508** | **NA** | N | 1 | 2 | 1 | 2 | PTV | | p.Arg286* | | 1/1 | NR | NR | 1/1 |
| **30093711** | **1** | N | 2 | 2 | 2 | 2 | PTV | | p.Arg78Glyfs*2 | 2 (1) | NR | NR | 1/1 | 0/1 |
| **30093711** | **10** | N | 1 | 2 | 1 | 2 | PTV | | p.Arg239* | 2 (1) | 1/1 | 1/1 | 1/1 | 0/1 |
| **30093711** | **14** | N | 1 | 2 | 1 | 2 | PTV | | p.Arg286* | 2 (1) | 1/1 | 1/1 | 1/1 | 1/1 |
| **30093711** | **15** | Y | 5 | 6 | 5 | 6 | PTV | | p.Asn315Argfs*4 | 8.5 (4) | 0/5 | 0/2 | 0/2 | 0/4 |
| **30093711** | **19** | Y | 3 | 4 | 3 | 4 | PTV | | p.Arg422* | 4.1 (2) | 1/3 | 1/1 | 0/1 | 1/3 |
| **30093711** | **20** | N | 2 | 2 | 2 | 2 | PTV | | p.Asn437Metfs*21 | 7.3 (2) | 1/2 | 0/2 | 0/2 | 0/2 |
| **30093711** | **21** | N | 1 | 2 | 1 | 2 | PTV | | p.Asn437Metfs*21 | 0.16 (1) | 1/1 | 1/1 | 1/1 | 1/1 |
| **30093711** | **23** | N | 1 | 2 | 1 | 2 | PTV | | p.Phe467Leufs*51 | 0.2 (1) | 1/1 | 0/1 | 0/1 | 1/1 |
| **30093711** | **26** | Y | 3 | 3 | 3 | 3 | PTV | | p.Arg555* | 11 (2) | 2/3 | 0/3 | 0/3 | 0/3 |
| **30093711** | **27** | N | 1 | 3 | 1 | 2 | PTV | | p.Arg555* | 2 (1) | 0/1 | 1/1 | 1/1 | 0/1 |
| **30093711** | **28** | N | 1 | 2 | 1 | 2 | PTV | | p.Arg615Serfs*47 | 8 (1) | 1/1 | 0/1 | 0/1 | 1/1 |
| **30093711** | **29** | Y | 2 | 2 | 2 | 2 | PTV | | p.Arg838* | 12.5 (2) | 0/1 | 0/2 | 0/1 | 0/2 |

| | | | | | | | | | | | | | | |
|---|---|---|---|---|---|---|---|---|---|---|---|---|---|---|
| **30093711** | **3** | N | 2 | 2 | 2 | 2 | PTV | c.279+1G >A | | 6 (1) | 1/1 | 0/1 | 0/1 | 1/1 |
| **30093711** | **31** | N | 1 | 2 | 1 | 2 | PTV | | p.Arg838* | 7 (1) | 0/1 | 1/1 | 1/1 | 0/1 |
| **30093711** | **33** | N | 2 | 2 | 2 | 2 | PTV | | p.Arg843* | 6.5 (2) | 1/2 | 0/2 | 1/2 | 0/2 |
| **30093711** | **34** | N | 1 | 2 | 1 | 2 | PTV | | p.Arg874* | 7 (1) | 0/1 | 0/1 | 1/1 | 0/1 |
| **30093711** | **35** | N | 1 | 2 | 1 | 2 | PTV | | p.Arg874* | 0.6 (1) | 0/1 | 0/1 | 1/1 | 0/1 |
| **30093711** | **37** | N | 1 | 2 | 1 | 2 | Other variant (Missense) | | p.Trp905Cys | 7 (1) | 1/1 | 1/1 | 1/1 | 0/1 |
| **30093711** | **38** | N | 1 | 2 | 1 | 2 | PTV | | p.Tyr920* | 1 (1) | 1/1 | 1/1 | 1/1 | 0/1 |
| **30093711** | **39** | N | 1 | 2 | 1 | 2 | Other variant (Missense) | | p.Ala928Val | 14 (1) | 0/1 | 1/1 | 1/1 | 0/1 |
| **30093711** | **40** | N | 1 | 2 | 1 | 2 | PTV | | p.Ala951Profs *38 | 2 (1) | 1/1 | 1/1 | 0/1 | 0/1 |
| **30093711** | **41** | N | 1 | 2 | 1 | 2 | Other variant (Missense) | | p.Arg997Cys | 0.6 (1) | 1/1 | 0/1 | 1/1 | 0/1 |
| **30093711** | **42** | N | 1 | 2 | 1 | 2 | PTV | c.3021+1 G>A | | 0 (1) | 1/1 | 1/1 | 1/1 | 1/1 |
| **30093711** | **44** | N | 1 | 2 | 1 | 2 | PTV | | p.Ala1077Asp fs*82 | 2 (1) | 0/1 | 0/1 | 0/1 | 0/1 |
| **30093711** | **45** | N | 2 | 2 | 2 | 2 | PTV | | p.Arg1087* | 10 (1) | 1/2 | 0/2 | NR | 0/2 |
| **30093711** | **46** | N | 1 | 2 | 1 | 2 | PTV | c.3330+5 G>C | | 4 (1) | 0/1 | 1/1 | 1/1 | 0/1 |
| **30093711** | **47** | N | 1 | 2 | 1 | 2 | Other variant (Missense) | | p.Ser1169Arg | 6 (1) | 0/1 | 0/1 | 0/1 | 0/1 |
| **30093711** | **49** | N | 1 | 2 | 1 | 2 | PTV | | p.Val1211_Leu1214delinsIleHisLeuHis | 8 (1) | 1/1 | 0/1 | 0/1 | 0/1 |
| **30093711** | **5** | N | 1 | 2 | 1 | 2 | PTV | | p.Gln107* | 4 (1) | 1/1 | 0/1 | 1/1 | 0/1 |
| **30093711** | **52** | N | 1 | 2 | 1 | 2 | PTV | | p.Arg1332* | 11 (1) | 1/1 | 1/1 | 0/1 | 1/1 |
| **30093711** | **53** | N | 1 | 2 | 1 | 2 | PTV | | p.Leu1344* | 1.1 (1) | 1/1 | 1/1 | 0/1 | NR |
| **30093711** | **54** | Y | 4 | 5 | 4 | 5 | PTV | | p.Glu1385* | 12 (4) | 1/4 | 2/4 | 3/4 | 0/4 |
| **30093711** | **55** | N | 1 | 2 | 1 | 2 | Other variant (Missense) | | p.Ala1392Val | 4 (1) | 0/1 | 0/1 | 1/1 | 0/1 |
| **30093711** | **56** | Y | 2 | 3 | 2 | 3 | PTV | | p.Glu174Lysfs *100 | 12 (2) | 2/2 | 0/2 | 1/2 | 0/2 |
| **30093711** | **59** | N | 2 | 2 | 2 | 2 | PTV | Deletion exons 31-42 | | 13 (1) | 0/1 | 0/2 | 0/1 | 0/1 |
| **30093711** | **6** | N | 1 | 2 | 1 | 2 | PTV | | p.Tyr127Ilefs* 51 | 5 (1) | 1/1 | 0/1 | 0/1 | NR |
| **30093711** | **61** | N | 1 | 2 | 1 | 2 | PTV | Deletion exons 8-19 | | 7 (1) | 0/1 | 0/1 | 1/1 | 0/1 |
| **30093711** | **7** | N | 1 | 2 | 1 | 2 | PTV | | p.Gly142Trpfs *3 | 0.6 (1) | 0/1 | 1/1 | 1/1 | 0/1 |
| **30093711** | **9** | N | 1 | 2 | 1 | 2 | Other variant (Missense) | | p.Met181Lys | 0.09 (1) | 1/1 | 0/1 | 0/1 | 0/1 |

| | | | | | | | | | | | | | | |
|---|---|---|---|---|---|---|---|---|---|---|---|---|---|---|
| **30767899** | **NA** | Y | 3 | 3 | 3 | 3 | Other variant (Missense) | | p.Leu1573Pro | 8 (1) | 0/3 | 0/3 | 1/1 | NR |
| **30868116** | **2** | N | 1 | 2 | 1 | 2 | PTV | c.4427-2A>G | | 0.75 (1) | 1/1 | NR | NR | 0/1 |
| **31225799** | **NA** | Y | 7 | 9 | 6 | 8 | PTV | c.280-1 G>A | | 0.25 (3) | 3/9 | 1/3 | NR | NR |
| **32848577** | **A** | N | 1 | 2 | 1 | 2 | PTV | | p.Val151Serfs *27 | 4 | 0/1 | 0/1 | 0/1 | 0/1 |
| **32848577** | **C** | Y | 2 | 3 | 2 | 3 | PTV | | p.Arg838* | 11.5 (2) | 0/2 | 0/2 | 0/2 | 0/2 |
| **32848577** | **E** | N | 1 | 2 | 1 | 2 | Other variant (Missense) | | p.Tyr7Cys | 11 (1) | 0/1 | 0/1 | 0/1 | 0/1 |
| **32848577** | **F** | Y | 2 | 3 | 2 | 3 | Other variant (Missense) | | p.Tyr836Cys | 2 (2) | 1/2 | 0/2 | 0/2 | 0/2 |
| **32848577** | **G** | Y | 2 | 3 | 2 | 3 | Other variant (Missense) | | p.Pro1031His | 5.5 (2) | 1/2 | 0/2 | 0/2 | 0/2 |
| **32848577** | **H** | N | 1 | 2 | 1 | 2 | Other variant (Missense) | | p.Pro1031His | 1.75 (2) | 0/1 | 0/1 | 0/1 | 0/1 |
| **32848577** | **I** | N | 1 | 2 | 1 | 2 | Other variant (Missense) | | p.Pro1031His | 1.8 (2) | 0/1 | NR | 0/1 | 0/1 |
| **32848577** | **J** | N | 1 | 2 | 1 | 2 | Other variant (Missense) | | p.Pro1031His | 2 (2) | 0/1 | 0/1 | 0/1 | 0/1 |
| **32848577** | **L** | N | 1 | 2 | 1 | 2 | Other variant (Missense) | | p.Gly1545Ser | 3 (1) | 0/1 | 0/1 | 0/1 | 0/1 |
| **33461085** | **2** | N | 2 | 2 | 2 | 2 | PTV | | p.Gly765Alafs *29 | 9.52 (2) | 1/2 | 1/2 | 0/1 | 1/2 |
| **33461085** | **3** | N | 2 | 2 | 2 | 2 | Other variant (Missense) | | p.Leu948Pro | 5 (2) | 1/2 | 1/2 | 1/1 | 1/2 |
| **34239491** | **B/2** | N | 1 | 4 | 1 | 4 | Other variant (Missense) | c.1729G>A | p.Val555Ile | | 0/1 | 0/1 | NR | NR |
| **34239491** | **C/3** | N | 1 | 2 | 1 | 2 | PTV | c.515_516 delinsT | | 2 (1) | 0/1 | 0/1 | NR | NR |
| **34239491** | **D/4** | Y | 3 | 3 | 3 | 3 | Other variant (Missense) | c.3260G>A | p.Arg1087Glu | 9 (3) | 1/3 | 0/3 | NR | NR |
| **35177946** | **3** | N | 1 | 2 | 1 | 2 | PTV | c.1287+1 G>A | | 59 (1) | 1/1 | 0/1 | 1/1 | 0/1 |
| **35907814** | **NA** | N | 1 | 2 | 1 | 2 | PTV | | p.Gln566fs | 0.4 (1) | 0/1 | 0/1 | NR | 0/1 |
| **36604176** | **1** | Y | 4 | 5 | 4 | 5 | PTV | c.364_368 del | p.Val122Hisfs *20 | 10.5 (4) | 1/4 | 1/4 | 1/4 | 0/4 |
| **36604176** | **2** | Y | 4 | 8 | 4 | 8 | PTV | c.461_462 del | p.Gly154Valfs *4 | 10 (5) | 2/4 | 0/4 | 1/4 | 0/4 |
| **36604176** | **3** | Y | 3 | 3 | 3 | 3 | PTV | c.1264C>T | p.Arg422* | 8 (5) | 0/4 | 0/4 | 0/3 | 0/3 |
| **37259768** | **I** | N | 1 | 2 | 1 | 2 | PTV | c.85_86insTGTTCCCTCACATCAAGCTTGG | | 0.25 (1) | 1/1 | 1/1 | NR | 1/1 |
| **37259768** | **II** | N | 2 | 2 | 2 | 2 | PTV | c.1342G>T | p.Glu448* | 9.5 (1) | 1/1 | 1/1 | NR | 0/1 |

| | | | | | | | | | | | | | | |
|---|---|---|---|---|---|---|---|---|---|---|---|---|---|---|
| **37259768** | **13** | N | 1 | 2 | 1 | 2 | PTV | c.1663C>T | p.Arg555* | 0 (1) | 0/1 | 0/1 | NR | 0/1 |
| **37259768** | **14** | N | 1 | 2 | 1 | 2 | PTV | c.1696C>T | p.Gln566* | 9.67 (1) | 1/1 | 0/1 | NR | 0/1 |
| **37259768** | **15** | N | 1 | 2 | 1 | 2 | PTV | c.1699C>T | p.Arg567* | 9 (1) | 1/1 | 1/1 | NR | 1/1 |
| **37259768** | **16** | N | 1 | 2 | 1 | 2 | Other variant (Missense) | c.1814G>A | p.Arg605Gln | 0.25 (1) | 1/1 | 1/1 | NR | 0/1 |
| **37259768** | **17** | N | 1 | 2 | 1 | 2 | PTV | c.2236_2237del | p.Ser746fs | 0.04 (1) | 1/1 | 1/1 | NR | 1/1 |
| **37259768** | **18** | N | 1 | 2 | 1 | 2 | PTV | c.2452A>T | p.Lys818* | 3 (1) | 0/1 | 0/1 | NR | 0/1 |
| **37259768** | **19** | N | 1 | 2 | 1 | 2 | PTV | c.2512C>T | p.Arg838* | 0.008 (1) | 1/1 | 1/1 | NR | 1/1 |
| **37259768** | **21** | N | 1 | 2 | 1 | 2 | PTV | c.2664C>A | p.Tyr888fs | 0.167 (1) | 1/1 | 1/1 | NR | 1/1 |
| **37259768** | **22** | N | 1 | 2 | 1 | 2 | PTV | c.2750_2751delAG | p.Lys917fs | 1.33 (1) | 1/1 | 1/1 | NR | 0/1 |
| **37259768** | **23** | N | 1 | 2 | 1 | 2 | Other variant (Missense) | c.2771A>G | p.Tyr924Cys | 0.25 (1) | 1/1 | 1/1 | NR | 0/1 |
| **37259768** | **24** | N | 1 | 2 | 1 | 2 | Other variant (Missense) | c.2785G>A | p.Gly929Ser | 2.25 (1) | 1/1 | 1/1 | NR | 0/1 |
| **37259768** | **25** | N | 1 | 2 | 1 | 2 | Other variant (Missense) | c.2948T>C | p.Leu983Pro | 0.67 (1) | 1/1 | 1/1 | NR | 1/1 |
| **37259768** | **26** | N | 1 | 2 | 1 | 2 | Other variant (Missense) | c.2965T>C | p.Phe989Leu | 0.33 (1) | 1/1 | 1/1 | NR | 1/1 |
| **37259768** | **27** | N | 1 | 2 | 1 | 2 | PTV | c.3046C>T | p.Gln1016* | 0 (1) | 1/1 | 1/1 | NR | 0/1 |
| **37259768** | **28** | N | 1 | 2 | 1 | 2 | PTV | Deletion exon 34 | | 0.05 | 1/1 | 1/1 | NR | 1/1 |
| **37259768** | **29** | N | 1 | 2 | 1 | 2 | PTV | c.3696+1G>T | | 0.02 | 1/1 | 1/1 | NR | 1/1 |
| **37259768** | **3** | N | 2 | 2 | 2 | 2 | PTV | c.313A>T | p.Lys105* | 0.08 | 1/1 | 0/1 | NR | 0/1 |
| **37259768** | **30** | N | 2 | 2 | 2 | 2 | PTV | c.3814C>T | p.Gln1272* | 0.167 (1) | 1/1 | 0/1 | NR | 0/1 |
| **37259768** | **31** | N | 1 | 2 | 1 | 2 | PTV | c.3817C>T | p.Gln1273* | 0.167 (1) | 1/1 | 1/1 | NR | 0/1 |
| **37259768** | **32** | N | 1 | 2 | 1 | 2 | PTV | c.3872-3891del | p.Arg1291fs | 5.17 (1) | 1/1 | 0/1 | NR | 1/1 |

| | | | | | | | | | | | | | | |
|---|---|---|---|---|---|---|---|---|---|---|---|---|---|---|
| **37259768** | **33** | N | 1 | 2 | 1 | 2 | PTV | c.3994C>T | p.R1332* | 0.08 (1) | 1/1 | 0/1 | NR | 1/1 |
| **37259768** | **35** | N | 1 | 2 | 1 | 2 | Other variant (Missense) | c.4337G>A | p.Ser1446Asn | 1.42 (1) | 1/1 | 0/1 | NR | 0/1 |
| **37259768** | **36** | N | 1 | 2 | 1 | 2 | PTV | c.4360G>T | p.Glu1454* | 1.08 (1) | 0/1 | 0/1 | NR | 0/1 |
| **37259768** | **37** | N | 1 | 2 | 1 | 2 | PTV | c.4416delC | p.Phe1472fs | 5.5 (1) | 1/1 | 0/1 | NR | 1/1 |
| **37259768** | **4** | N | 1 | 2 | 1 | 2 | PTV | c.363+4A>G | | 0.167 (1) | 1/1 | 1/1 | NR | 1/1 |
| **37259768** | **5** | N | 1 | 2 | 1 | 2 | PTV | c.414-2A>T | | 7.25 (1) | 0/1 | 0/1 | NR | 0/1 |
| **37259768** | **6** | N | 1 | 2 | 1 | 2 | Other variant (Missense) | c.533G>A | p.Ser178Asn | 0.167 (1) | 1/1 | 0/1 | NR | 1/1 |
| **37259768** | **7** | N | 1 | 2 | 1 | 2 | Other variant (Missense) | c.536G>T | p.Cys179Phe | 1.08 (1) | 1/1 | 0/1 | NR | 0/1 |
| **37259768** | **9** | N | 2 | 2 | 2 | 2 | PTV | c.727C>T | p.Arg243* | 0.33 (1) | 1/1 | 1/1 | NR | 1/1 |
| **37263295** | **NA** | N | 1 | 3 | 1 | 3 | Other variant (Missense) | c.2763A>T | p.Leu921Phe | 7 (1) | 1/1 | 1/1 | NR | 0/1 |
| **37422919** | **3** | Y | 3 | 4 | 3 | 4 | PTV | Deletion exons 12–17 | | 0.75 (3) | 1/3 | 1/3 | 1/3 | 0/3 |
| **38261030** | **NA** | N | 1 | 2 | 1 | 2 | PTV | c.2799G>A | p.Trp933* | 0.33 (1) | 1/1 | NR | NR | 1/1 |
| **38752894** | **2** | N | 1 | 2 | 1 | 2 | PTV | c.194-1G>C | | 1.4 (1) | 1/1 | 0/1 | NR | 0/1 |
| **38752894** | **3** | Y | 3 | 4 | 3 | 4 | PTV | c.414-1G>T | | 3.6 (1) | 1/3 | 1/3 | NR | 0/2 |
| **38752894** | **4** | Y | 2 | 3 | 2 | 3 | PTV | c.990delG | p.Gln331Argfs*5 | 0.7 (1) | 0/1 | 0/2 | NR | 0/1 |
| **38752894** | **5** | Y | 3 | 4 | 3 | 4 | PTV | c.1018delG | p.Val340Trpfs*11 | 5.7 (1) | 0/2 | 0/3 | NR | 0/2 |
| **38752894** | **6** | N | 1 | 2 | 1 | 2 | PTV | c.1416_1417insC | p.Ser473Profs*44 | 3 (1) | 1/1 | 1/1 | NR | 0/1 |
| **38752894** | **7** | N | 1 | 2 | 1 | 2 | PTV | c.1453C>T | p.Arg485* | 10.8 (1) | 0/1 | 0/1 | NR | 0/1 |
| **38752894** | **8** | N | 1 | 2 | 1 | 2 | PTV | c.1696delC | p.Gln566fs | 0.4 (1) | 0/1 | 0/1 | NR | 0/1 |
| **38974383** | **NA** | Y | 2 | 4 | 2 | 4 | PTV | c.1217+2T>A | | | 1/1 | 1/1 | NR | 0/2 |

[a]From available individual data

NA: not available, Y: yes, N: no, PTV: protein truncating variant, DR: drug resistant, ID: intellectual disability, NR: not reported

#### Supplementary Table 3. Additional sources of data.

| PMID | Family ID | Additional clinical information from different source |
|---|---|---|
| **23542697** | **H** | PMID 27066554, Family E |
| **23542697** | **A1** | PMID 24585383, Family B and PMID 9851433 |
| **23542701** | **S** | PMID 10825362, Family S and PMID 25623524, Family 1 |
| **23542701** | **L** | PMID 10825362, Family L |
| **23542701** | **Q** | PMID 10825362, Family Q |
| **23542701** | **N** | PMID 10825362, Family N |
| **23542701** | **O** | PMID 10825362, Family O |
| **24585383** | **C** | PMID 23542697, Family I |
| **26000329** | **NA** | PMID 24502525, Family 1 |
| **26505888** | **18** | PMID 29125946 |
| **26505888** | **23** | PMID 37003255 |
| **31225799** | **NA** | PMID 34239491, Family A/1 |

NA: not available
